# Highly accurate and sensitive diagnostic detection of SARS-CoV-2 by digital PCR

**DOI:** 10.1101/2020.03.14.20036129

**Authors:** Lianhua Dong, Junbo Zhou, Chunyan Niu, Quanyi Wang, Yang Pan, Sitong Sheng, Xia Wang, Yongzhuo Zhang, Jiayi Yang, Manqing Liu, Yang Zhao, Xiaoying Zhang, Tao Zhu, Tao Peng, Jie Xie, Yunhua Gao, Di Wang, Yun Zhao, Xinhua Dai, Xiang Fang

## Abstract

The outbreak of COVID-19 caused by a novel Coronavirus (termed SARS-CoV-2) has spread to over 140 countries around the world. Currently, reverse transcription quantitative qPCR (RT-qPCR) is used as the gold standard for diagnostics of SARS-CoV-2. However, the positive rate of RT-qPCR assay of pharyngeal swab samples are reported to vary from 30∼60%. More accurate and sensitive methods are urgently needed to support the quality assurance of the RT-qPCR or as an alternative diagnostic approach.

**METHODS:** We established a reverse transcription digital PCR (RT-dPCR) protocol to detect SARS-CoV-2 on 194 clinical pharyngeal swab samples, including 103 suspected patients, 75 close contacts and 16 supposed convalescents.

**RESULTS:** The limit of blanks (LoBs) of the RT-dPCR assays were ∼1.6, ∼1.6 and ∼0.8 copies/reaction for ORF 1ab, N and E genes, respectively. The limit of detection (LoD) was 2 copies/reaction. For the 103 fever suspected patients, the sensitivity of SARS-CoV-2 detection was significantly improved from 28.2% by RT-qPCR to 87.4% by RT-dPCR. For close contacts, the suspect rate was greatly decreased from 21% down to 1%. The overall sensitivity, specificity and diagnostic accuracy of RT-dPCR were 90%, 100% and 93 %, respectively. In addition, quantification of the viral load for convalescents by RT-dPCR showed that a longer observation period was needed in the hospital for elderly patients.

**CONCLUSION:** RT-dPCR could be a confirmatory method for suspected patients diagnosed by RT-qPCR. Furthermore, RT-dPCR was more sensitive and suitable for low viral load specimens from the both patients under isolation and those under observation who may not be exhibiting clinical symptoms.

## Introduction

In late December 2019, a number cases of pneumonia infection were reported in Wuhan, Hubei Province, China. It was officially named as Coronavirus disease (COVID-19) by the World Health Organization (WHO) and has since spread to over 120 countries around the world *(1, 2)*. The pathogen causing the outbreak of disease was identified as a novel Coronavirus (termed SARS-CoV-2), belonging to the family *Coronaviridae*, order *Nidovirales*, all of which are enveloped, non-segmented positive-sense RNA viruses *(3, 4)*. According to the WHO and Chinese Center for Disease Control and Prevention (CDC), the current gold standard for the diagnosis of SARS-CoV-2 infection is based on the reverse transcript quantitative PCR (RT-qPCR).

However, RT-qPCR is reported to have issues of low positive rates for throat swab samples *(5)* and there were 3% of patients who had negative RT-qPCR test results at initial presentation while chest CT checks indicated typical symptoms of viral pneumonia*(6)*. In order to identify and hospitalize COVID-19 patients in time, more sensitive and accurate tests are required.

Digital PCR (dPCR) is a technology which partitions nucleic acid molecules across a large number of smaller reactions and acquires amplification data of each partition at end point based on the intensity of fluorescence *(7-9)*. Quantification is performed by applying Poisson statistics to the proportion of the negative partitions to account for positive partitions that initially contained more than one target molecule. dPCR can offer greater precision than qPCR and is far simpler to use for copy number quantification due the binary nature in which the partitions are counted as positive or negative. Additionally, dPCR is more tolerant of PCR inhibition compared with qPCR due to partitioning and because it is an end-point PCR measurement and consequently less dependent on high PCR efficiency *(10, 11)*.

In this study, we established one step RT-dPCR protocol for detection of ORF1ab open reading frame 1ab (ORF1ab) and nucleocapsid protein (N) and E gene of SARS-CoV-2. Moreover, we compared RT-qPCR and RT-dPCR on 194 clinical samples and found RT-dPCR can significantly improve the sensitivity and diagnostic accuracy of Coronavirus disease (COVID-19).

## Materials and Methods

### ETHICS STATEMENT

Data collection of cases and close contacts were determined by the National Health Commission of the People’s Republic of China to be part of a continuing public health outbreak investigation and were thus considered exempt from institutional review board approval. The analysis was performed on existing samples collected during standard diagnostic tests, posing no extra burden to patients.

### STUDY DESIGN

103 febrile suspected SARS-CoV-2 infected patients, 75 close contacts and 16 supposed convalescents were chosen in this study. Positive, negative and suspect results were all included in the chosen specimens according to the RT-qPCR test.

### CLINICAL SAMPLES

Respiratory samples were obtained during February and March 2020 from patients hospitalized or close contacts tested by Beijing CDC (BJCDC), Wuhan CDC (WHCDC) and a government designated clinical test laboratory (Wuhan considerin laboratory for medical test, KXR). Samples were stored in viral transport medium (Yocon Biology) at 4□. RNA was extracted from clinical specimens within 24 hours by using the MagMAX-96 viral RNA isolation kit (Thermo Fisher Scientific). RNA extracts containing human coronaviruses (HCoV)-229E and (HCoV)-OC43 and avian influenza virus RNAs A/H3N2 and A/H1N1 Virus and Influenza B/Victoria Virus were provided by BJCDC. Extracted RNA was stored at -80□.

### ONE STEP REVERSE TRANSCRIPTION DPCR (RT-DPCR)

The primer and probe sequences for detecting N and ORF1ab gene target of the SARS-CoV-2 published by Chinese CDC were used for this study*(12)*. For detecting E gene target, primer and probe recommended by world health organization (WHO) was used*(13)*. The 20 μL reaction mixture comprises5 μL of Supermix, 2 μL of reverse transcriptase, 1 μL of 300 mM DTT from One-Step RT-ddPCR Advanced Kit for Probes (Cat. #1864021, Bio-Rad), 1 μL of mixture of primers and probe and 11 μL of RNA template. Each reaction mix was converted to droplets using the QX200 Droplet Generator (Bio-Rad,), transferred to a 96-well plate (Cat. #120019285, Bio-Rad), heat sealed and amplified in a GeneAmp System 9700 thermal cycler (Applied Biosystems). The thermal cycling conditions were as follows: 45°C for 10 min (reverse transcription); 95°C for 5 min; and 40 cycles of 95°C for 15 sec, and 58°C for 30 sec. The cycled plate was then transferred to the QX200 Droplet Reader (Bio-Rad) and analyzed using the QuantaSoft droplet reader software (V1.7.4, Bio-Rad). Reactions containing more than 10,000 droplets were treated as effective and involved in data analysis.

### LIMIT OF BLANK (LOB) AND DETECTION (LOD) OF RT-DPCR

To establish the limit of blank (LoB) *(14)*, 60 blank measurements were obtained from 3 blank samples on three days. 70 to 76 measurements from 4-5 samples with low concentration (1 to 3 cp/reaction) were used to determine the limit of detection (LoD) according to the CLSI guideline of EP17-A*(15)*.

### RT-QPCR

Three different commercial RT-qPCR kits (H&R from Shanghai Huirui Biotechnology Co., Ltd, BioGerm from Shanghai BioGerm Medical Biotechnology and Daan from Daan Gene Co., Ltd) were used for the detection. Briefly, a 25-μL reaction containing 7.5 μL of PCR reaction buffer, 5 µL of primer and probe mixture and 5∼11 μL of RNA was prepared. Thermal cycling was performed at 50 °C for 15 min for reverse transcription, followed by 95°C for 5 min and then 45 cycles of 95 °C for 10 s, 55 °C for 45 s in ABI 7500 RT-PCR thermocycler. Data analysis was performed using software of ABI 7500 RT-PCR thermocycler.

## Results

### DYNAMIC RANGE OF THE RT-DPCR ASSAY

The linear range was investigated by varying the mean copy number per droplet, denoted as λ*(16)*. The precision or relative error of RT-dPCR is related to λ because of RT-dPCR relies on the Poisson statistics to account for droplets with multiple molecules*(17)*. The upper limit of the linearity was 7.8 copies/partition tested by N gene assay. To determine the lower limit of all three assays, serial dilutions of each RNA transcript were prepared (Supplemental Table 1). The measured targets matched the anticipated values in each tested interval. A good linearity (0.93<slope<1.02, R^2^ ≥ 0.9997) between the measured RNA target and the prepared value was observed over the range from approximately 10^4^ to 10^0^ copies/reaction (Fig. 1). Reactions containing a mean of 60 E, 66 N or 11 ORF1ab copies fulfilled the criterion for an LoQ with a CV lower than 25%.

**Table 1.** Results of SARS-CoV-2 by RT-qPCR and RT-dPCR on 103 febrile suspected patients.

| Patient number | Reported result | ORF1ab gene |  | N gene |  | E gene | Result by RT-dPCR | clinical status |
| --- | --- | --- | --- | --- | --- | --- | --- | --- |
|  |  | RT-qPCR | RT-dPCR | RT-qPCR | RT-dPCR | RT-dPCR |  |  |
|  |  | Ct | copies/reaction | Ct | copies/reaction | copies/reaction |  |  |
| P1 | Positive | 31.71 | 7248 | 27.9 | 4892 | 5156.4 | Positive | Viral pneumonia, Fever cough |
| P2 | Positive | 32.06 | 3208 | 29.51 | 2212 | 3160 | Positive | Viral pneumonia, Fever cough |
| P3 | Positive | 32.95 | 2460 | 30.79 | 1064 | 1480.6 | Positive | Viral pneumonia, Fever cough |
| P4 | Positive | 34.18 | 1108 | 30.91 | 1048 | 840.8 | Positive | Viral pneumonia, Fever cough |
| P5 | Positive | 34.3 | 1062 | 33.06 | 242 | 842 | Positive | Viral pneumonia, Fever cough |
| P6 | Positive | 34.47 | 1046 | 33.86 | 226 | 836.4 | Positive | Viral pneumonia, Fever cough |
| P7 | Positive | 35.07 | 542 | 33.99 | 228.1 | 428.6 | Positive | Viral pneumonia, Fever cough |
| P8 | Suspect | 35.6 | 302 | 32.14 | 562 | 424.4 | Positive | Viral pneumonia, Fever cough |
| P9 | Suspect | 36.18 | 187.6 | 35.89 | 58 | 130.2 | Positive | Viral pneumonia, Fever cough |
| P10 | Suspect | 36.19 | 179.8 | 32.97 | 426 | 132 | Positive | Viral pneumonia, Fever cough |
| P11 | Suspect | 36.1 | 186 | 35.49 | 24 | 70 | Positive | Viral pneumonia, Fever cough |
| P12 | Suspect | 36.58 | 170 | 34.81 | 11 | 34 | Positive | Viral pneumonia, Fever cough |
| P13 | Suspect | 36.87 | 110 | 35.27 | 1.8 | 24 | Positive | Viral pneumonia, Fever cough |
| P14 | Suspect | 36.91 | 96 | 35.02 | 9.4 | 28 | Positive | Viral pneumonia, Fever cough |
| P15 | Suspect | 36.99 | 102 | 36.57 | 8.6 | 9 | Positive | Viral pneumonia, Fever cough |
| P16 | Suspect | 37.91 | 84 | 37.41 | 24.6 | 68.8 | Positive | Viral pneumonia, Fever cough |
| P17 | Positive | 38.29 | 76 | 34.59 | 204.8 | 120.4 | Positive | Viral pneumonia, Fever cough |
| P18 | Suspect | 38.29 | 74.6 | 0 | 66 | 86 | Positive | Viral pneumonia, Fever cough |
| P19 | Suspect | 38.38 | 72 | 33.18 | 214.8 | 104 | Positive | Viral pneumonia, Fever cough |
| P20 | Negative | 39.41 | 32 | 0 | 54 | 98 | Positive | Viral pneumonia, Fever cough |
| P21 | Suspect | 39.88 | 28.6 | 34.04 | 186 | 92.4 | Positive | Viral pneumonia, Fever cough |
| P22 | Negative | 39.95 | 27 | 45 | 57 | 89 | Positive | Viral pneumonia, Fever cough |
| P23 | Suspect | 39.97 | 24.2 | 36.68 | 23.6 | 90.4 | Positive | Viral pneumonia, Fever cough |
| P24 | Suspect | 40.22 | 8.8 | 38.98 | 12.4 | 16.7 | Positive | Viral pneumonia, Fever cough |
| P25 | Negative | 40.66 | 6.4 | 45 | 4.2 | 8.4 | Positive | Viral pneumonia, Fever cough |
| P26 | Suspect | 41.2 | 7.2 | 32.84 | 18.4 | 14.8 | Positive | Viral pneumonia, Fever cough |
| P27 | Negative | 41.39 | 4.6 | 39.8 | 5.8 | 10.8 | Positive | Viral pneumonia, Fever cough |
| P28 | Suspect | 41.91 | 3.4 | 37.46 | 13.4 | 6.4 | Positive | Viral pneumonia, Fever cough |
| P29 | Negative | 42.45 | 3.4 | 43.63 | 3.2 | 5.4 | Positive | Viral pneumonia, Fever cough |
| P30 | Negative | 43.28 | 4.8 | 0 | 2.4 | 5.8 | Positive | Viral pneumonia, Fever cough |
| P31 | Negative | 43.28 | 5.4 | 0 | 0 | 4.9 | Positive | Viral pneumonia, Fever cough |
| P32 | Suspect | 43.51 | 3.8 | 0 | 2.2 | 2.4 | Positive | Viral pneumonia, Fever cough |
| P33 | Negative | 43.98 | 2.6 | 0 | 4.8 | 42.4 | Positive | Viral pneumonia, Fever cough |
| P34 | Suspect | 43.99 | 2.6 | 37.09 | 3.2 | 5.8 | Positive | Viral pneumonia, Fever cough |
| P35 | Suspect | 37.16 | 94 | 36.07 | 0 | 20 | Positive | Viral pneumonia, Fever cough |
| P36 | Suspect | 37.21 | 70 | 34.51 | 11.4 | 38 | Positive | Viral pneumonia, Fever cough |
| P37 | Suspect | 37.29 | 58 | 34.04 | 3.2 | 32 | Positive | Viral pneumonia, Fever cough |
| P38 | Suspect | 37.47 | 48 | 36.42 | 50 | 44 | Positive | Viral pneumonia, Fever cough |
| P39 | Suspect | 37.94 | 52 | 35.74 | 6.4 | 30 | Positive | Viral pneumonia, Fever cough |
| P40 | Suspect | 38.29 | 32 | 35.44 | 6.4 | 24 | Positive | Viral pneumonia, Fever cough |
| P41 | Suspect | 38.305 | 32 | 36.884 | 0 | 13.4 | Positive | Viral pneumonia, Fever cough |
| P42 | Suspect | 38.83 | 40 | 38.63 | 0 | 20 | Positive | Viral pneumonia, Fever cough |
| P43 | Suspect | 38.95 | 34 | 40 | 3.8 | 0 | Positive | Viral pneumonia, Fever cough |
| P44 | Suspect | 39.01 | 26 | 38.56 | 0 | 11.8 | Positive | Viral pneumonia, Fever cough |
| P45 | Suspect | 39.11 | 34 | 37.13 | 0 | 24 | Positive | Viral pneumonia, Fever cough |
| P46 | Suspect | 39.81 | 20 | 37.01 | 3.6 | 0 | Positive | Viral pneumonia, Fever cough |
| P47 | Suspect | 39.9 | 12.4 | 36.56 | 0 | 20 | Positive | Viral pneumonia, Fever cough |
| P48 | Suspect | 39.94 | 16 | 36.82 | 2 | 12 | Positive | Viral pneumonia, Fever cough |
| P49 | Suspect | 40 | 0 | 38.68 | 0 | 0 | Negative | Viral pneumonia, Fever cough |
| P50 | Negative | 40 | 0 | 39.68 | 0 | 0 | Negative | Viral pneumonia, Fever cough |
| P51 | Suspect | 40 | 10 | 37.88 | 0 | 12 | Positive | Viral pneumonia, Fever cough |
| P52 | Suspect | 40 | 8.6 | 38.748 | 0 | 1.8 | Positive | Viral pneumonia, Fever cough |
| P53 | Suspect | 40.42 | 3.4 | 39.12 | 0 | 0 | Positive | Viral pneumonia, Fever cough |
| P54 | Negative | 40.47 | 0 | 40 | 3 | 1.6 | Positive | Viral pneumonia, Fever cough |
| P55 | Negative | 40.65 | 3 | 40 | 2.2 | 0 | Positive | Viral pneumonia, Fever cough |
| P56 | Suspect | 40.65 | 1.8 | 38.41 | 0 | 0 | Suspect | Viral pneumonia, Fever cough |
| P57 | Negative | 40.97 | 1.8 | 40 | 0 | 0 | Suspect | Viral pneumonia, Fever cough |
| P58 | Suspect | 41.02 | 0 | 38.89 | 0 | 6 | Positive | Viral pneumonia, Fever cough |
| P59 | Negative | 41.2 | 0 | 40 | 1.8 | 0 | Suspect | Viral pneumonia, Fever cough |
| P60 | Negative | 42.5 | 12 | 40 | 0 | 0 | Positive | Viral pneumonia, Fever cough |
| P61 | Suspect | NA | 1.2 | 38.08 | 0 | 0 | Suspect | Viral pneumonia, Fever cough |
| P62 | Negative | ND* | 0 | 39.84 | 2.4 | 1.4 | Positive | Viral pneumonia, Fever cough |
| P63 | Negative | ND | 0 | 40.14 | 1.8 | 2.6 | Positive | Viral pneumonia, Fever cough |
| P64 | Negative | ND | 0 | 39.71 | 0 | 0 | Negative | Viral pneumonia, Fever cough |
| P65 | Negative | ND | 2.1 | 39.43 | 2.2 | 3.4 | Positive | Viral pneumonia, Fever cough |
| P66 | Suspect | ND | 0 | 37.96 | 1.6 | 2.2 | Positive | Viral pneumonia, Fever cough |
| P67 | Negative | ND | 0 | 39.86 | 0 | 0 | Negative | Viral pneumonia, Fever cough |
| P68 | Suspect | ND | 1.4 | 38.1 | 0 | 2.4 | Positive | Viral pneumonia, Fever cough |
| P69 | Suspect | ND | 0 | 39 | 0 | 2 | Positive | Viral pneumonia, Fever cough |
| P70 | Suspect | ND | 0 | 36.89 | 2 | 0 | Positive | Viral pneumonia, Fever cough |
| P71 | Negative | ND | 0 | 39.29 | 2.2 | 1.4 | Positive | Viral pneumonia, Fever cough |
| P72 | Suspect | ND | 0 | 32.23 | 2.6 | 0 | Positive | Viral pneumonia, Fever cough |
| P73 | Suspect | ND | 0 | 35.76 | 2.4 | 4.6 | Positive | Viral pneumonia, Fever cough |
| P74 | Negative | ND | 0 | 40.24 | 2.8 | 3 | Positive | Viral pneumonia, Fever cough |
| P75 | Suspect | ND | 0 | 39.44 | 1 | 2.8 | Positive | Viral pneumonia, Fever cough |
| P76 | Suspect | ND | 0 | 38.27 | 0 | 0 | Negative | Viral pneumonia, Fever cough |
| P77 | Suspect | ND | 0 | 37.36 | 2.8 | 2.4 | Positive | Viral pneumonia, Fever cough |
| P78 | Negative | ND | 0 | 42.09 | 0 | 0 | Negative | Viral pneumonia, Fever cough |
| P79 | Negative | ND | 0 | 40.98 | 1.8 | 1.8 | Suspect | Viral pneumonia, Fever cough |
| P80 | Suspect | ND | 0 | 39.39 | ND | 1.4 | Suspect | Viral pneumonia, Fever cough |
| P81 | Negative | ND | 0 | 40.88 | ND | 1.4 | Suspect | Viral pneumonia, Fever cough |
| P82 | Positive | 39.15 | 7.2 | 38.27 | ND | 4.4 | Positive | Viral pneumonia, Fever cough |
| P83 | Negative | ND | 0 | ND | 0 | 5.2 | Positive | Viral pneumonia, Fever cough |
| P84 | positive | 25.44 | 3440 | 26.32 | 4700 | 880 | Positive | Viral pneumonia, Fever cough |
| P85 | positive | 29.41 | 36 | 28.23 | 242 | 3.8 | Positive | Viral pneumonia, Fever cough |
| P86 | positive | 29.45 | 16 | 29.11 | 328 | 12 | Positive | Viral pneumonia, Fever cough |
| P87 | positive | 37.62 | 0 | 31.72 | 12 | 12 | Positive | Viral pneumonia, Fever cough |
| P88 | positive | 36.45 | 5.2 | 34.28 | 0 | 24 | Positive | Viral pneumonia, Fever cough |
| P89 | positive | Negative | 12 | positive | - | 80 | Positive | Viral pneumonia, Fever cough |
| P90 | positive | positive | 3940 | positive | - | 31500 | Positive | Viral pneumonia, Fever cough |
| P91 | positive | positive | 4540 | positive | - | 29600 | Positive | Viral pneumonia, Fever cough |
| P92 | positive | Negative | 4 | positive | - | 34 | Positive | Viral pneumonia, Fever cough |
| P93 | positive | Negative | 0 | positive | - | 6 | Positive | Viral pneumonia, Fever cough |
| P94 | positive | Negative | 12 | positive | - | 42 | Positive | Viral pneumonia, Fever cough |
| P95 | positive | positive | 142 | positive | - | 3.4 | Positive | Viral pneumonia, Fever cough |
| P96 | positive | positive | 22 | positive | - | 98 | Positive | Viral pneumonia, Fever cough |
| P97 | positive | Negative | 2 | positive | - | 22 | Positive | Viral pneumonia, Fever cough |
| P98 | positive | Negative | 4.4 | positive | - | 13.6 | Positive | Viral pneumonia, Fever cough |
| P99 | positive | Negative | 1.8 | positive | - | 14.2 | Positive | Viral pneumonia, Fever cough |
| P100 | positive | Negative | 0 | positive | - | 4.8 | Positive | Viral pneumonia, Fever cough |
| P101 | positive | Negative | 2 | positive | - | 8.2 | Positive | Viral pneumonia, Fever cough |
| P102 | positive | Negative | 5.8 | positive | - | 8.2 | Positive | Viral pneumonia, Fever cough |
| P103 | positive | Negative | 3.4 | positive | - | 12.6 | Positive | Viral pneumonia, Fever cough |

ND\*, Ct Not detectable.

**Fig 1.**
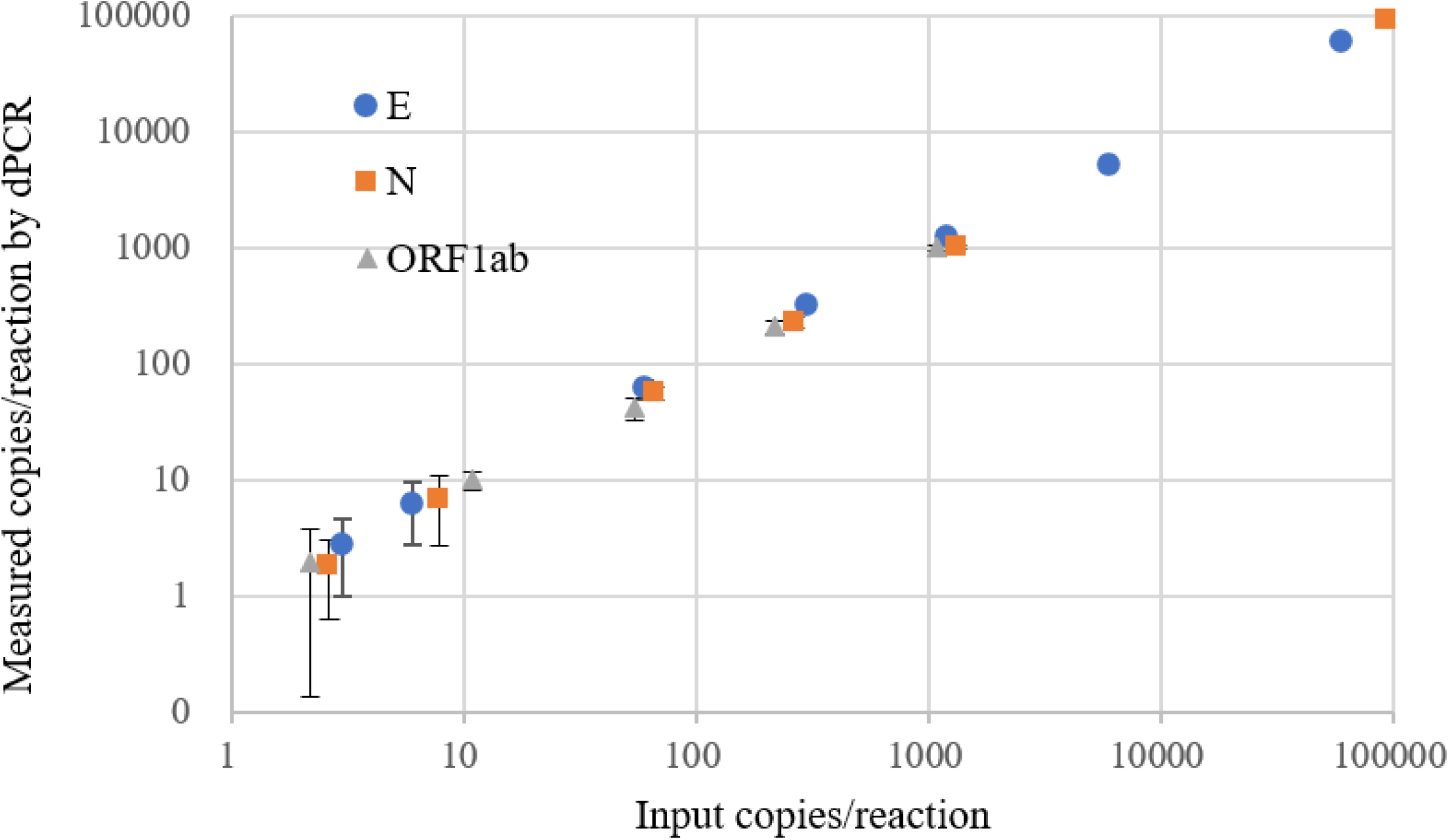
Validated range of the RT-dPCR assays for E, ORF1ab and N gene. Evaluation of linearity of samples containing E, ORF1ab and N gene molecules over the extended λ range (0.0002 <λ< 7.83). Data are shown in mean with standard deviation for each dilution series (3≤n≤10).

### ESTABLISHMENT OF LOB AND LOD FOR RT-DPCR ASSAY

Sixty blank measurements obtained from 3 blank samples were analyzed to determine the LoB. As the distribution of the 60 blank measurements is skewed (Supplemental Fig. 1), the LoB was estimated nonparametrically as the 95th percentile of the measurements. The 15 highest blank values for each target are displayed in Supplemental Table 2. The 95th percentile corresponds to the 57.5 ordered observation (=60*(0.95/60+0.5))*(15)*. Linear interpolation between the 57th and 58th observation yields a LoB estimate of 1.6, 1.6, and 0.8 copies/reaction for E, ORF1ab and N, respectively.

**Table 2.** Comparison of RT-qPCR and RT-dPCR on SARS-CoV-2 RNA measurement of close contacts.

| Patient number | Reported result | ORF1ab gene |  | N gene |  | E gene | Result by dPCR | clinical status |
| --- | --- | --- | --- | --- | --- | --- | --- | --- |
|  |  | qPCR | dPCR | qPCR | dPCR | dPCR |  |  |
|  |  | Ct | copies/reaction | Ct | copies/reaction | copies/reaction |  |  |
| P1 | Suspect | 0 | 2.2 | 37.37 | 0 | 7 | Positive | Isolated observation, Asymptomatic |
| P2 | Positive | 38.1 | 16 | 36.76 | 0 | 3.2 | Positive | Isolated observation, Asymptomatic |
| P3 | Suspect | 0 | 2.2 | 37.93 | 0 | 0 | Positive | Isolated observation, Asymptomatic |
| P4 | Suspect | 37.26 | 9.6 | 0 | 5.3 | 3 | Positive | Isolated observation, Asymptomatic |
| P5 | Positive | 42.63 | 3.6 | 38.39 | 0 | 8.6 | Positive | Isolated observation, lower fever, cough |
| P6 | Positive | 39.9 | 1.8 | 38.41 | 0 | 5 | Positive | Isolated observation, lower fever, cough |
| P7 | Suspect | 0 | 1.4 | 36.7 | 1.4 | 1.6 | Suspect | Isolated observation, Asymptomatic |
| P8 | Suspect | 0 | 1.6 | 36.84 | 0 | 3 | Positive | Isolated observation, Asymptomatic |
| P9 | Suspect | 0 | 0 | 36.58 | 0 | 7 | Positive | Isolated observation, Asymptomatic |
| P10 | Suspect | 38.21 | 1.8 | 0 | 2.7 | 0 | Positive | Isolated observation, Asymptomatic |
| P11 | Suspect | 35.31 | 2.2 | 0 | 0 | 1.4 | Positive | Isolated observation, Asymptomatic |
| P12 | Suspect | 36.45 | 3.4 | 0 | 0 | 0 | Positive | Isolated observation, Asymptomatic |
| P13 | positive | 35.72 | 16.6 | 34.41 | 9.8 | 15.6 | Positive | Isolated observation, Asymptomatic |
| P14 | Suspect | 0 | 98 | 36.75 | 0 | 14 | Positive | Isolated observation, Asymptomatic |
| P15 | Suspect | 36.88 | 20 | 0 | 0 | 3 | Positive | Isolated observation, Asymptomatic |
| P16 | positive | 37.53 | 26 | 36.11 | 3.2 | 16 | Positive | Isolated observation, Asymptomatic |
| P17 | positive | 25.38 | 1682 | 22.8 | 4700 | 5640 | Positive | Isolated observation, Asymptomatic |
| P18 | Suspect | 0 | 22 | 35.47 | 0 | 3 | Suspect | Isolated observation, Asymptomatic |
| P19 | Suspect | 39.74 | 5 | 0 | 0 | 12 | Positive | Isolated observation, Asymptomatic |
| P20 | Suspect | 0 | 28 | 39.21 | 1.6 | 94 | Positive | Isolated observation, Asymptomatic |
| P21 | Negative | >40 | 8 | 0 | 0 | 8 | Positive | Isolated observation, Asymptomatic |
| P22 | positive | 31.78 | 214 | 30.56 | 72 | 104 | Positive | Isolated observation, Asymptomatic |
| P23 | Suspect | 0 | 900 | 35.37 | 0 | 32 | Positive | Isolated observation, Asymptomatic |
| P24 | Suspect | 35.32 | 26 | 0 | 0 | 28 | Positive | Isolated observation, Asymptomatic |
| P25 | positive | 33.12 | 28 | 33.65 | 38 | 14.2 | Positive | Isolated observation, Asymptomatic |
| P26 | positive | 24.5 | 5280 | 26.8 | 3980 | 2650 | Positive | Isolated observation, Asymptomatic |
| P27 | positive | 33.31 | 64 | 32.41 | 74 | 50 | Positive | Isolated observation, Asymptomatic |

For determining the LoD of ORF1ab gene assay, 76 measurements were performed on five samples in 3 different runs on three different days to ensure the total assay variation is reflected. The distribution of the 76 measurement results from low concentration samples is not Gaussian (Supplemental Fig. 2A) and so that nonparametric statistics was used according to the guideline of EP17-A. Consequently, the LoD is determined to be 2 copies/reaction, the lowest level material where the β-percentile is 5 %.

**Fig 2.**
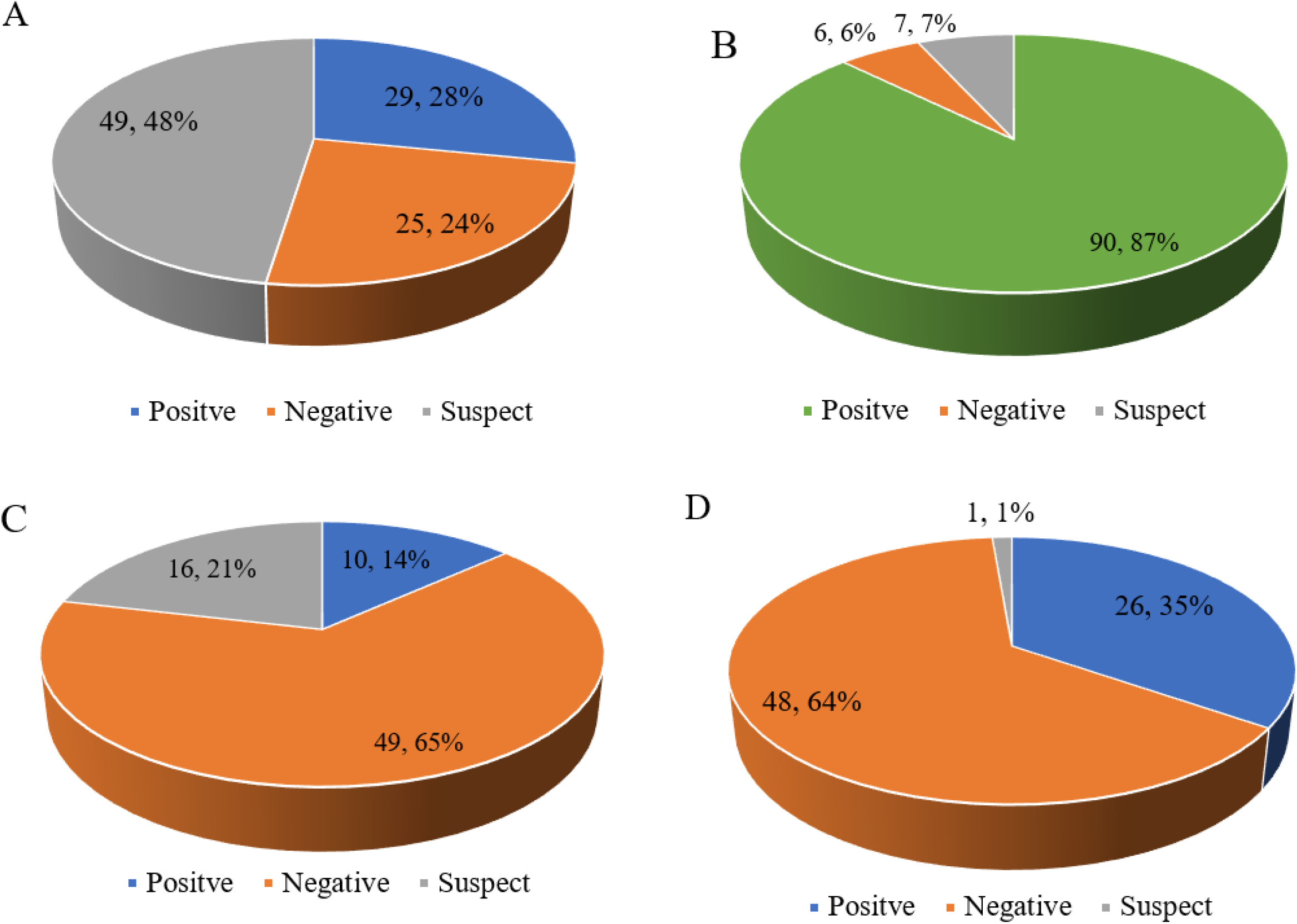
Diagnostics of SARS-CoV-2 by RT-qPCR and RT-dPCR on 103 febrile suspected patients and 75 close contacts. 25 samples positive, 29 negative and 49 suspected were reported by RT-qPCR (A). 90 positive, 6 negative and 7 equivocal were determined by RT-dPCR (B).10 positive, 49 negative and 16 suspected were reported by RT-qPCR (C). 26 positive, 48 negative and 1 equivocal were determined by dPCR (D).

To determine the LoD of N and E assay, 83 measurements of E assay on 5 samples and 71 measurements of N assay on 4 samples were performed in 4 different runs. Similar to ORF1ab gene, the distribution of the 71 measurements for N gene and 83 measurements are not Gaussian (Supplemental Fig. 2B and 2C), and so that nonparametric statistics was used. Consequently, the LoD is determined to be 2 copies/ reaction.

### SPECIFICITY TESTING

The Specificity of the assays for ORF1ab and E gene has been tested in a previous report *(13)*. To further validate the specificity of all assays, Influenza virus and other human coronavirus were collected. All assays were tested on human clinical nucleic acid samples at National institute of Metrology, China. All tests returned negative results (see Supplemental Table 3).

**Table 3.**
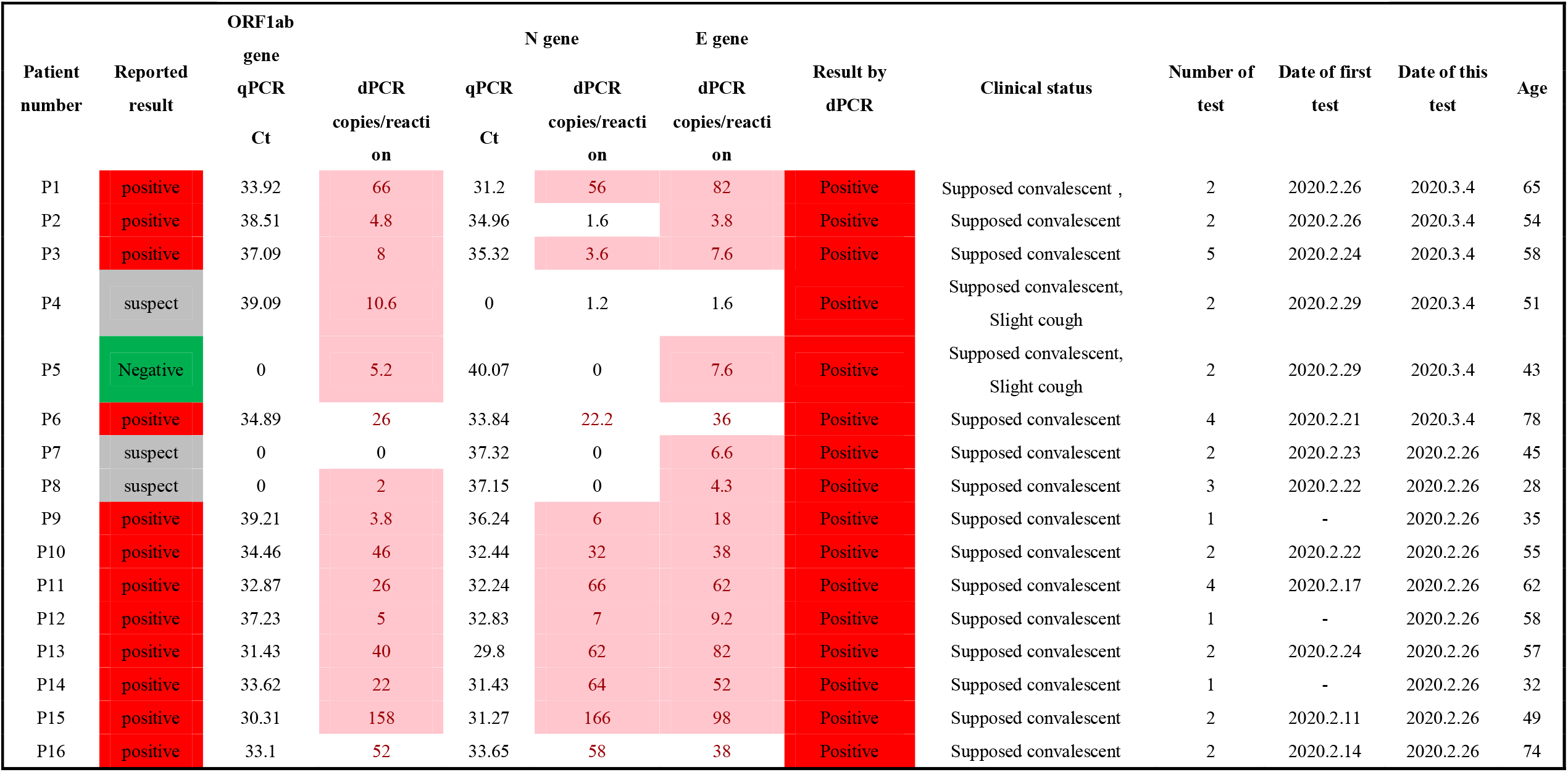
Comparison of SARS-CoV-2 RNA measurement on convalescent patients by RT-qPCR and RT-dPCR.

### COMPARISON BETWEEN RT-QPCR AND RT-RT-DPCR ON FEBRILE SUSPECTED PATIENTS

103 pharyngeal swabs were collected from febrile suspected SARS-CoV-2 infected patients and the relevant information is listed in Table 1. Among the 103 specimens, 81 (P1 to P81) were tested at KXR with the H&R qPCR kit and 7 (P82-88) were tested at WHCDC by the Daan qPCR kit. Firstly, the criteria claimed by the H&R kit manufacturer are: Ct value≤35 are positive, Ct value >39.2 are negative, and 35<Ct<39.2 are equivocal. The criteria of the Daan qPCR kit are: Ct >40, negative, Ct ≤40, positive, and equivocal if only one gene with Ct ≤ 40 and no amplification for another gene. According to such criteria, 14 positive, 25 negative and 49 suspected SARS-CoV-2 infections were determined and this was reported to the clinical doctors. For RT-dPCR, three targets are tested in parallel and the determination of a positive result should meet the following criteria: quantification of any one of the three gene targets is ≥2 copies/ reaction. If no positive droplet was detected in FAM channel but positive droplets were detected in VIC indicating RNAseP positive for human reference control*(11)*, the sample can be judged negative. If 0 < result < 2, it should be attributed to equivocal and need further check. According to such criteria, 44 out of 49 suspects and 17 out of 25 negatives were corrected to be positive by RT-dPCR and the positive rate significantly increased. No positive droplet was detected for the 6 negatives and copy numbers were quantified under the established LoD for 7 suspects infections, due to either no virus sampled or ultra-low viral load in these specimens.

15 samples (from P89-P103) were tested at BJCDC with BioGerm qPCR kit and assays recommended by Chinese CDC. Ct values were not available and only negative or positive information were reported. Single gene target positive was determined to be SARS-CoV-2 positive based on parallel test with a commercial kit and Chinese CDC assays. Therefore, these 15 samples were reported positive by BJCDC. 8 qPCR negatives for ORF1ab were positive tested by RT-dPCR, showing high sensitivity for ORF1ab by RT-dPCR. Only 3 negatives for ORF1ab which can be complemented by E gene targets.

Among the 103 specimens, 29 positive, 25 negative and 49 suspected were reported by RT-qPCR. However, 61 samples including 17 negative and 44 suspected tested by qPCR were confirmed to be positive by RT-dPCR, thus 90 patients in total whose SARS-CoV-2 nucleic acid were positive tested can be diagnosed with COVID-19. All the 103 patients were confirmed SARS-CoV-2 infection according to a follow-up survey. The sensitivity of SARS-CoV-2 detection was significantly improved from 28.2% to 87.4% for the 103 patients (Fig. 2A and 2B).

### COMPARISON ON CLOSE CONTACTS AND CONVALESCENT

75 specimens were collected from contacts and close contacts. 48 specimens from contacts were reported negative based RT-qPCR test by BJCDC on Feb 6 and were confirmed by RT-dPCR on Feb 7 in Supplemental Table 4. According to a follow-up survey, all of them were in good health and isolation was lifted after 14 days.

27 specimens (Table 2 and Fig. 2C and 2D) were detected at WHCDC by RT-qPCR with a kit from Daan gene on March 2, 4 and 6. According to RT-qPCR result, 10 positive, one negative and 16 suspect were reported. It is very difficult to detect the SARS-CoV-2 nucleic acids due the low viral load at the early stage for the close contacts. However, 15 out of 16 equivocal and one negative can be determined positive by RT-dPCR. The suspect rate was significantly decreased from 21% down to 1% according to the detection of RT-dPCR. Consequently, except 6 patients can not be tracked, the rest 10 RT-dPCR positive were confirmed as SARS-CoV-2 infected patients based on a follow-up survey.

Furthermore, among the 16 specimens corrected by RT-dPCR, 6 persons (P14,18-21and P23 in Supplemental Table 5) were directed for secondary testing following an initial negative test 2 to 10 days prior. Based on RT-qPCR results, further isolation and observation was still needed to be conducted as the testing result is suspect or negative and no clinical symptoms were observed for them. However, if based on RT-dPCR, all the six patients can be diagnosed with COVD-19 infected by SARS-CoV-2 and treatment could be conducted earlier. This indicates RT-dPCR is more sensitive and suitable for low viral load specimens from the patients under isolation and observation without clinical symptoms, which is in agreement with the very recent online report *(18)*.

Additionally, 16 pharyngeal swabs were collected from convalescent patients (Table 3). 12 positive, 3 suspect and 1 negative were reported by qPCR. However, all of these 16 patients are diagnosed to be positive by RT-dPCR, indicating that all of them still need to be observed in hospital. Regardless the sampling time, correlation between age and the RNA virus copy number was analyzed (Fig.3). Interestingly, except P15, with increasing age, the copy number of viral load was much higher, which indicates a longer observation in the hospital is needed. We set up the threshold of 15, 20 and 25 copies/reaction for ORF1ab, N and E, respectively. The ORF1ab, N and E gene copy number were higher than their threshold for 100% patients elder than 60 and 75% (6 out 8 patients) elder than 55 (the median).

**Fig 3.**
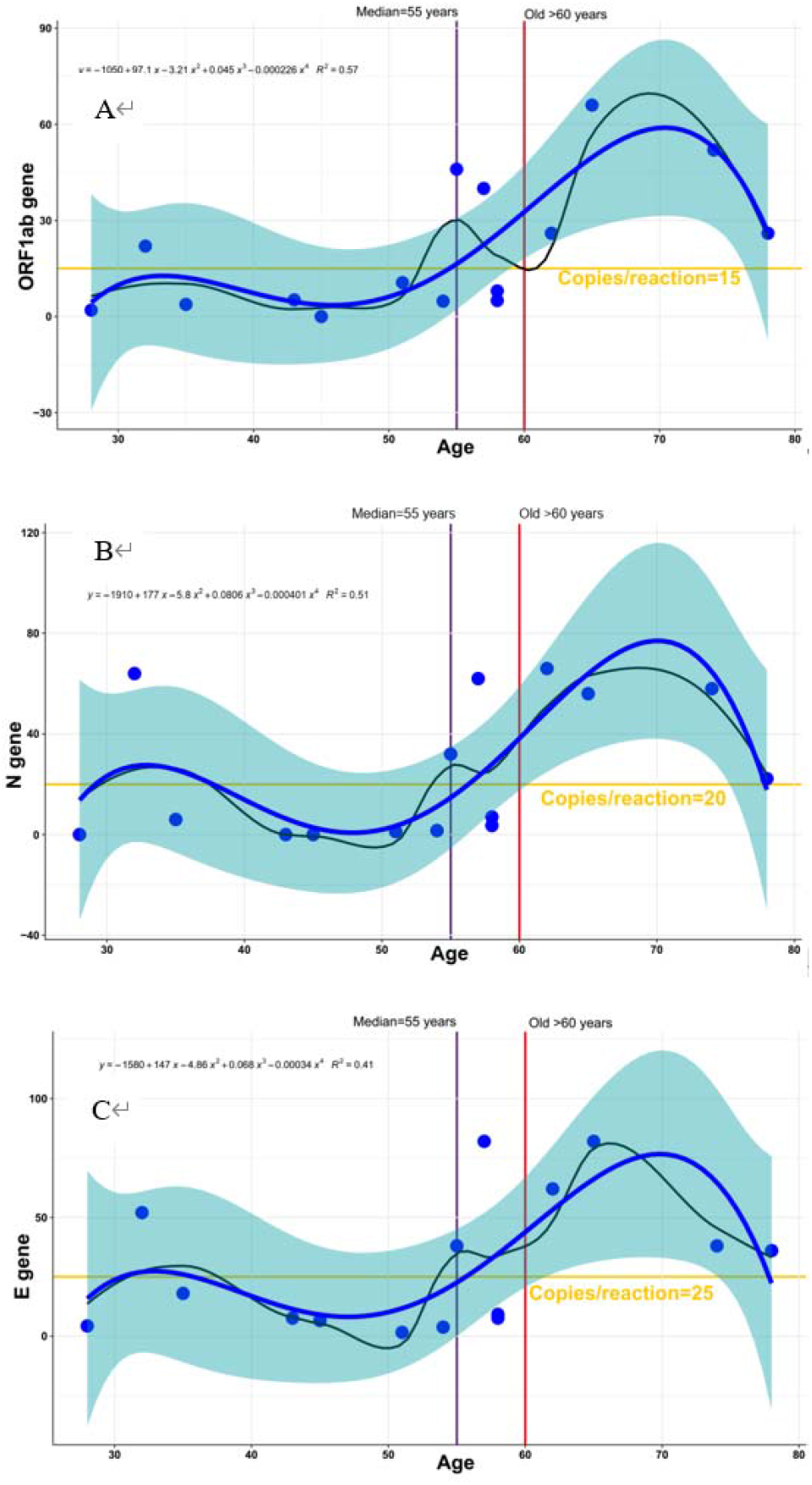
Scatter plot of copies/reaction values of 15 patients detected by RT-dPCR for ORF1ab (A), N (B) and E (C). The samples are arranged by ages (from younger to older). The blue curves fitted by lasso regression show the distribution of copies/reaction values. The shading that underlies the lasso curves denotes the 95% confidence intervals. The black curve represents the underlying relationship between copies/reaction values and ages of the convalescent patients, which was fitted by the methods of liner models. The liner model was labeled on the left-top of picture. The horizontal line represents the threshold of 15, 20 and 25 copies/reaction for ORF1ab, N and E, respectively. The ORF1ab, N and E gene copy number were higher than their threshold for 100% patients elder than 60 and 75% (6 out 8 patients) elder than 55 (the median).

## Discussion

RT-qPCR, as the standard method of diagnostics of SARS-CoV-2, plays an important role in this outbreak, though a low positive rate has been reported *(5)*. A number of factors could affect RT-PCR testing results including sample collection and transportation, RNA extraction and storage, and proper performance of the kit *(19)*. More recently, more than 145 RT-qPCR kits have been developed by the in vitro diagnostic manufactures (IVDs) in China *(20)*. Among the RT-qPCR kits, those with low sensitivity would cause high false negative rate or high equivocal rate. For the equivocal results it is necessary to conduct a retest and this would improve the positive rate of RT-qPCR. However, in the clinical practice under the current situation, it is impossible to do a same day retest due to the daily burden of thousands of incoming samples. The testing laboratory should initially report a result based on a single test, while secondary sampling for a later retest does not need to be sent to the same laboratory. Therefore, availability of a highly sensitive and accurate confirmatory method is of particular importance for the diagnosis of SARS-CoV-2 in this outbreak.

Currently, besides RT-qPCR, other methods such as next generation sequencing (NGS) and immunological detection of IgM and IgG could be used as confirmatory methods for diagnosis of COVID-19 according to the latest guideline of Diagnosis and Treatment of Pneumonitis Caused by SARS-CoV-2 (trial seventh version) published by National Health Commission *(21)*. This would decrease the false negative rate by applying multiple methods. However, diagnostics of nucleic acids is still considered as the gold standard as this is the most direct way to detect the presence of the virus. Thus, digital PCR method could be a powerful complement method because it can significantly improve the sensitivity for the suspect patients. The overall sensitivity and diagnostic accuracy of RT-qPCR in our study were 36% and 53%, respectively, according to our follow-up survey on the 188 samples(6 out of 194 can not be tracked). The RT-qPCR sensitivity is in agreement with the previous report for the throat swab samples *(5)*. However, both sensitivity and diagnostic accuracy of RT-dPCR dramatically increased to 90 % and 93%, respectively. Furthermore, it is very sensitive for the very low viral load in close contacts and suitable for monitoring the change of the viral load in the convalescent patients. An additional advantage of quantification of SARS-CoV-2 copy number by RT-dPCR is that comparisons can be conducted between different dates and different laboratories as absolute quantitation of targets by RT-dPCR provides high concordance between sites, runs and operators *(14, 22, 23)*. However, it is not possible to compare Ct values on different runs or different machines. Thus, RT-dPCR is an ideal method to for measuring the change of virus load in the convalescent patients.

This work demonstrates that RT-dPCR significantly improves accuracy and reduces the false negative rate of diagnostics of SARS-CoV-2, which could be a powerful complement to the current RT-qPCR. Furthermore, RT-dPCR is more sensitive and suitable for low virus load specimens from the patients under isolation and observation who may not be exhibiting clinical symptoms.

## Data Availability

All relevant data are available upon request.

## Supplemental Material

Supplemental material is available

### Author Contributions

All authors confirmed they have contributed to the intellectual content of this paper and have met the following 4 requirements: (a) significant contributions to the conception and design, acquisition of data, or analysis and interpretation of data; (b) drafting or revising the article for intellectual content; final approval of the published article; and (d) agreement to be accountable for all aspects of the article thus ensuring that questions related to the accuracy or integrity of any part of the article are appropriately investigated and resolved.

### Authors’ Disclosures or Potential Conflicts of Interest

Upon manuscript submission, all authors completed the author disclosure form. Disclosures and/or potential conflicts of interest:

### Employment or Leadership

X. Fang

### Consultant or Advisory Role

None declared.

### Stock Ownership

None declared.

### Honoraria

None declared.

### Research Funding

Fundamental Research Funds for Central Public welfare Scientific research Institutes sponsored by National Institute of Metrology, P.R. China (31-ZYZJ2001/AKYYJ2009).

### Expert Testimony

None declared.

### Patents

None declared.

## Acknowledgments

We would like to thank Academy of Military Medical Sciences for providing the purified virus RNA for method validation, Dr. Huggett from the National Measurement Laboratory, NML for his valuable comments and Bio-Rad Laboratories, China for donating the one step RT-RT-dPCR mastermix.

